# Evaluation of the Panbio™ COVID-19 Ag Rapid Test at an Emergency Room in a Hospital in São Paulo, Brazil

**DOI:** 10.1101/2021.03.15.21253313

**Authors:** Klinger Soares Faíco-Filho, Francisco Estivallet Finamor Júnior, Luiz Vinícius Leão Moreira, Paulo Ricardo Gessolo Lins, Alberto Fernando Oliveira Justo, Nancy Bellei

**Author notes:** Corresponding Author: Nancy Bellei, Laboratório de Virologia, Division of Infectious Diseases, Department of Medicine, Escola Paulista de Medicina (EPM), Rua Pedro de Toledo 781 -15 andar CEP- 04039 032, São Paulo, Brazil.

## Abstract

**Introduction:** The performance characteristics of the Panbio™ COVID-19 Ag test were evaluated against RT-PCR – considered the gold-standard for the detection of SARS-CoV-2 – at an emergency room in São Paulo, Brazil. The study aimed to determine the sensitivity, specificity, positive percent agreement (PPA), and negative percent agreement (NPA) as compared to RT-PCR.

**Methods:** Specimens from 127 suspected patients were tested by both the Panbio™ COVID-19 Ag test and by RT-PCR.

**Results:** Evaluation of the agreement between the Panbio™ COVID-19 Ag test and RT-PCR indicated an overall sensitivity of 84% (95%CI, 75-93.8%) and an overall specificity of 98% (95%CI, 96-98.8%). The Panbio™ COVID-19 Ag test showed 97% sensitivity and 99% NPA for subjects with Ct values ≤ 25 (N=37).

**Conclusion:** The Panbio™ COVID-19 Ag test is suitable for use as a diagnostic test for the rapid screening of patients presenting COVID-19 symptoms, or those suspected with being infected, prior to being admitted to hospital.

## INTRODUCTION

In late 2019, an outbreak of respiratory illness of unknown etiology was reported in Wuhan City, Hubei Province, China. The International Committee for Taxonomy of Viruses (ICTV) named the virus SARS-CoV- 2 and is the cause of the global COVID-19 pandemic which, as of 4^th^ March 2021, has claimed more than 2.5 million lives (https://covid19.who.int/). The preferred diagnostic test for COVID-19 ╌ nucleic acid amplification test (NAAT) using reverse transcriptase polymerase chain reaction (RT-PCR) ╌ uses fluid from the nasal or nasopharyngeal cavities and the time to get results can vary from same day to a week^1,2,3^. Alternatively, a COVID-19 antigen test using nasal or nasopharyngeal samples can produce results in minutes^3^. A positive antigen test result is considered accurate, but low viral load in the sample can produce false-negative results^5,6^.

The Abbott Panbio™ COVID-19 Ag Rapid Test device is an *in vitro* rapid diagnostic test intended to aid in the diagnosis of COVID-19. The Panbio™ COVID-19 Ag Rapid Test device is a lateral flow immunochromatographic test used for the qualitative detection of antigens to SARS-CoV-2 in human tissue fluids obtained from nasal or nasopharyngeal swabs. The Panbio™ COVID-19 Ag platform uses a cassette containing a lateral flow test strip and is intended for use by trained healthcare professionals in point of care and hospital settings. The product may be used in both laboratory and non-laboratory environments that meet the requirements specified in the product’s Instructions for Use (IFU). In this study, the performance of the Panbio™ COVID-19 Ag Rapid test device using fluids obtained from nasopharyngeal swabs was compared to the study site’s standard of care RT-PCR test. The Panbio™ COVID-19 Ag test can be used for rapid screening of patients presenting COVID-19 symptoms, or those having contact with infected persons, prior to being admitted to hospital.

## MATERIALS AND METHODS

### 1. Study Population

This prospective study was conducted at São Paulo Hospital in São Paulo, Brazil. We included patients (≥ 18 years), treated in the emergency room and hospitalized for at least 24 hours, and met one of the following criteria: (1) people with symptoms suspected to be related to SARS-CoV- 2 and/or contact with COVID-19 infected persons, (2) decompensation of underlying disease, or (3) suggestive tomographic alteration (ground glass).

### 2. Testing Scheme

Nasopharyngeal swab samples were simultaneously tested using the Panbio™ COVID-19 Ag test and by RT-PCR. The Panbio™ COVID-19 Ag test was performed according to the product IFU^7^. Results were obtained and interpreted at 15 minutes after test start according to the IFU. The RT-PCR testing was performed according to the IFU for the GeneFinder™ COVID-19 Plus RealAmp Kit (OSANG Healthcare Co., Ltd.) targeting the *RdRp* (RNA-dependent RNA Polymerase), *E* (envelope) and *N* (nucleocapsid) SARS-CoV-2 genes^9^. Samples that produced inconclusive GeneFinder™ test results were tested using a second RT-PCR test – Mobius XGEN MASTER COVID-19 test – targeting the *ORF1ab* and *N* SARS-CoV-2 genes, per the IFU^10^. Test results for both RT-PCR methods were available within 6-24 hours.

### 3. Reference Testing

For this study, the performance of the Panbio™ COVID-19 Ag test was evaluated against the results of the standard test (RT-PCR). For RT-PCR, RNA was isolated from the subject’s nasopharyngeal swab using the Quick-RNA Viral Kit (Zymo Research, Irvine, CA), according to the manufacturer’s protocol^8^. After extraction, the RNA was used immediately, and the remaining RNA was stored at −80°C. The Master Mixture was prepared by mixing 10 µL of COVID-19 Plus Reaction Mixture and 5 µL of COVID-19 Plus Probe Mixture per sample; a sufficient amount of Master Mixture was prepared for all the samples and controls that were tested. Then, 15 µL of the Master Mixture was transferred to a 96-well plate to which either (a) 5 µL RNA sample, (2) 5 µL negative control (DEPC-treated water), or (3) 5 µL positive control (DNA plasmids encoding the SARS-CoV-2 *RdRp, E*, and *N* genes, and human *RNase P* gene), were added. The plate was sealed and centrifuged at 2,000 rpm for 10 seconds. The thermal cycling conditions were as follows: (1) 1 cycle at 50°C for 20 min, (2) 1 cycle at 95°C for 5 min, and (3) 45 cycles at 95°C for 15 sec and 58°C for 60 sec.

In contrast to the GeneFinder™ kit, the Master Mixture in the Mobius™ kit was used directly without further preparation. The 96-well plate was prepared by adding 15 µL of the Master Mixture to the sample wells, followed by either (1) 5 µL of the subject’s RNA, (2) 5 µL negative control (DEPC-treated water), or (3) 5 µL positive control (a synthetic cDNA of the *ORF1ab* and *N* SRS-CoV-2 genes). In addition, 1 µL of an internal control was added to all the subjects’ RNA samples. It is important to highlight that the composition of the internal control is not described by the manufacturer. The plate was sealed and centrifuged, as described above. The thermal cycling conditions were as follows: (1) 1 cycle at 45°C for 15 min, (2) 1 cycle at 95°C for 2 min, and (3) 45 cycles at 95°C for 10 sec and 60°C for 50 sec.

### 4. Statistical Analysis

To assess differences between patient subgroups, continuous variables were expressed by mean (and standard deviation) or median (and interquartile range) values and compared using the Student t-test, ANOVA, Kruskal-Wallis test and/or the Mann-Whitney *U* test. Categorical variables were presented as proportions and compared using the Chi-square test and the Fisher exact test. The distribution of continuous variables was checked by the Kolmogov-Smirnov test.

#### Binary logistic regression - exclusion of the constant factor

A Receiver Operating Characteristic (ROC) curve with evaluation of Area Under the Curve (AUC) and 95% Confidence Interval (CI) - Adjustment for the lower Ct value, the higher prediction of the test. All analyses were performed using the SPSS 26 program. Differences were considered statistically significant at p values lower than 0.05.

## RESULTS

A total of 127 subjects were included. The median age was 63 years (22-69) and there were more males than females: 54.3% vs 45.7%. The characteristics of the study population are given in Table 1. The Panbio™ COVID-19 Ag testing summary is shown in Table 2.

**Table 1:** RT-PCR Results and Clinical Characteristics of Study Population.

|  | PCR Negative<br>(N=57) | PCR Positive<br>(N=70) | P | Total<br>(N=127) |
| --- | --- | --- | --- | --- |
| Mean of age (± SD) | 54.74 ± 19.25 | 64.31 ± 14.83 | 0.002 | 60 ± 17.5 |
| Male gender (%) | 29 (50.9%) | 40 (57.1%) | 0.481 | 69 (54.3%) |
| Time of Symptoms (days, mean) | 5 (3; 7) | 6 (4; 7) | 0.06 | 5 (4; 7) |
| Ct Value* (mean) | 40 (40; 40) | 25 (21.75; 28) | <0.001 | 32 (25; 40) |

**Table 2:** Panbio™ COVID-19 Ag Test Results and Clinical Characteristics of Study Population.

|  | Antigen Negative<br>(N=67) | Antigen Positive<br>(N=60) | P | Total<br>(N=127) |
| --- | --- | --- | --- | --- |
| Mean of age (± SD) | 55.94 ± 18.60 | 64.57 ± 15.19 | 0.005 | 60 ± 17.5 |
| Male gender (%) | 35 (52.2%) | 34 (56.7%) | 0.617 | 69 (54.3%) |
| Time of Symptoms (days, mean) | 5 (3; 7) | 6 (4; 7) | 0.178 | 5 (4; 7) |
| Ct Value* (mean) | 40 (40; 40) | 24.5 (21; 28) | <0.001 | 32 (25; 40) |

Table 3 provides details of the 10 samples that were negative by the Panbio™ COVID-19 Ag test but were positive by RT-PCR using the GeneFinder™ COVID-19 Plus RealAmp Kit. The Ct values of the samples ranged from 21-34 and were collected from subjects 2-11 days post onset of symptoms.

**Table 3:** Details of Discordant Samples.

| Sample | Ct Value | Days Post Onset of Symptoms |
| --- | --- | --- |
| 1 | 21 | 8 |
| 2 | 26 | 2 |
| 3 |  | 4 |
| 4 |  | 7 |
| 5 | 28 | 10 |
| 6 | 30 | 7 |
| 7 | 31 | 11 |
| 8 | 32 | 4 |
| 9 | 34 | 5 |

Table 4 shows the Sensitivity and Specificity as a function of Ct. The results indicate that the Panbio™ COVID-19 Ag test has high sensitivity (97%) for samples with Ct values ≤ 25, with an overall sensitivity of 84% when all samples that are positive by RT-PCR (Ct values < 40) are included in the analysis. Conversely, the specificity of the Panbio™ COVID-19 Ag test is lower (59%) for samples with Ct values ≤ 25 but increases to 98% when all samples that are negative by RT-PCR are included (Ct values < 40).

**Table 4:**
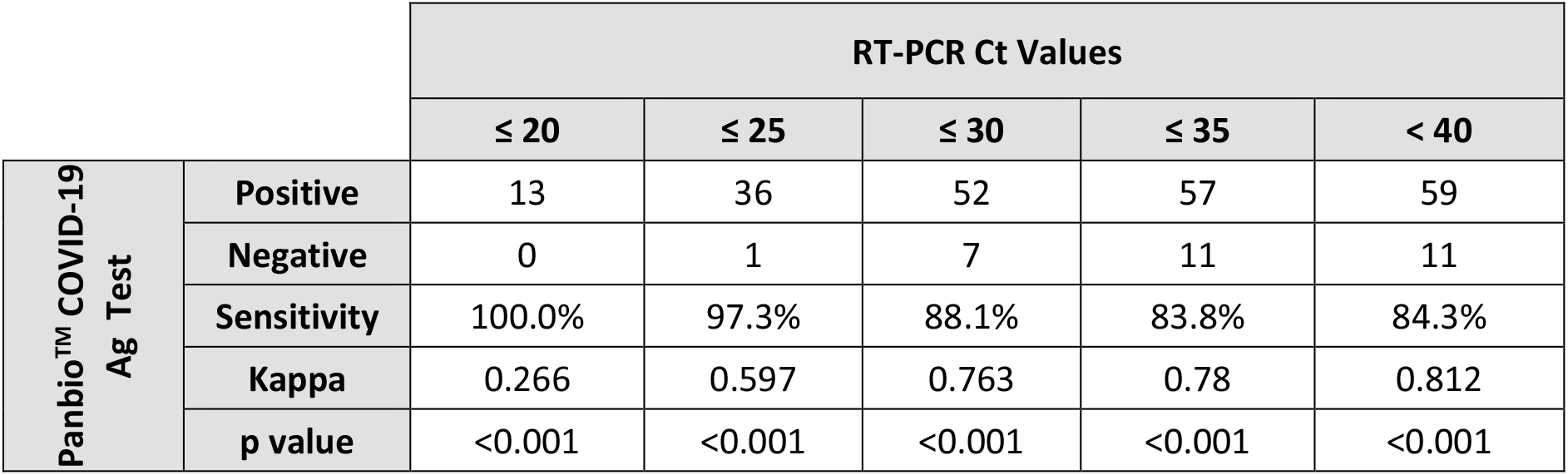
Sensitivity and Specificity as a Function of Ct.

Figure 1 shows the ROC curve of the Panbio™ COVID-19 Ag test results versus Ct values. The curve shows that the Panbio™ COVID-19 Ag test is highly sensitive. Prediction ROC curve for positive Antigen result based on the PCR CT value - AUC 0.949 (0.911 - 0.987) p <0.001.

**Figure 1:**
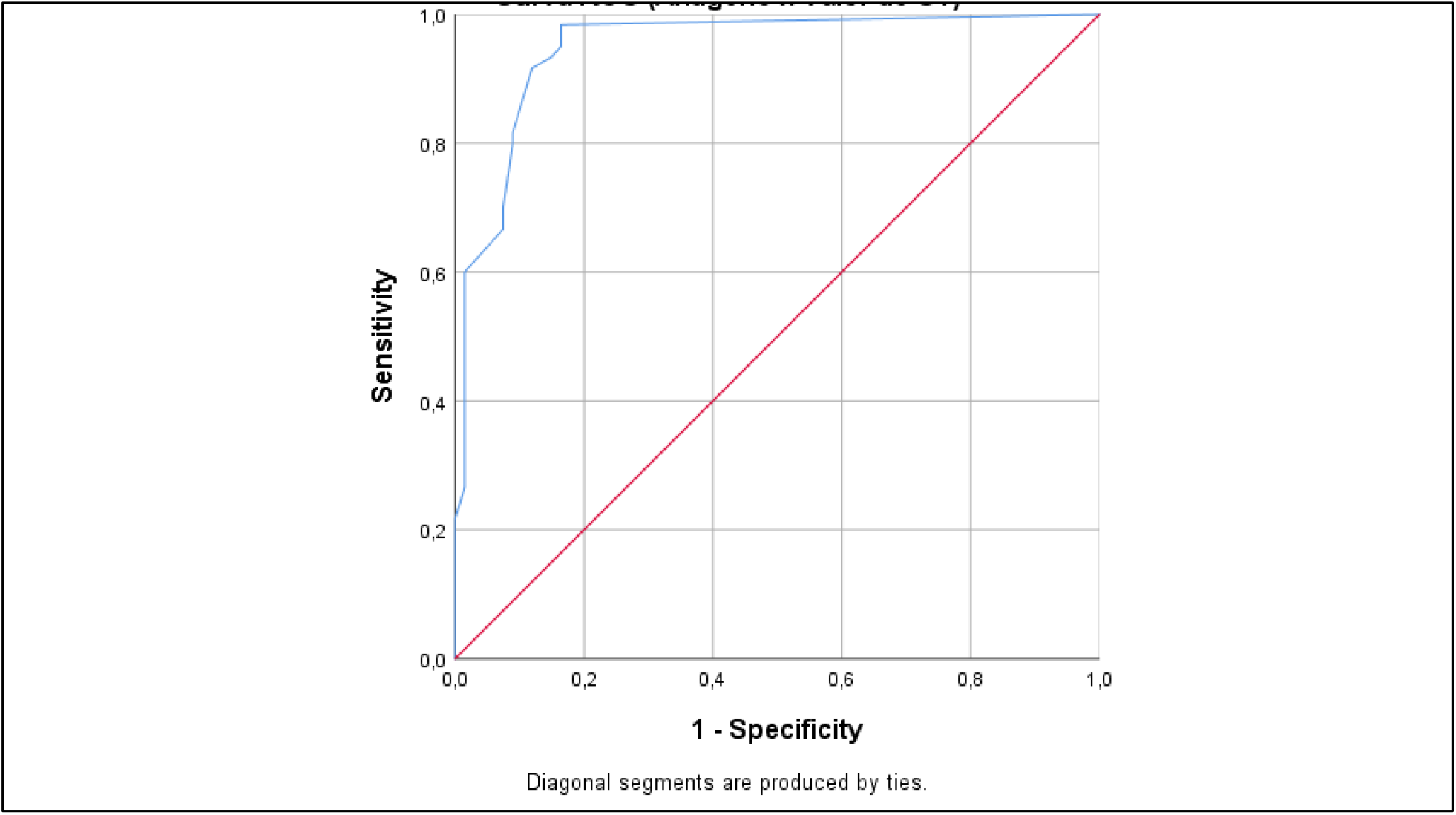
ROC Curve: Panbio™ COVID-19 Ag test results versus Ct.

Figure 2 is a graph that represents the distribution of Ct values over time for PCR-positive samples (N=70). The number of data points is less than 70 because some samples produced the same Ct values and had the same symptom day. No PCR-negative samples are presented here. The red squares represent PCR-positive samples that were tested as negative by the Panbio™ COVID-19 Ag test. We detected the highest viral loads soon after symptom onset, which then gradually decreased but viral loads were very heterogeneous.

**Figure 2:**
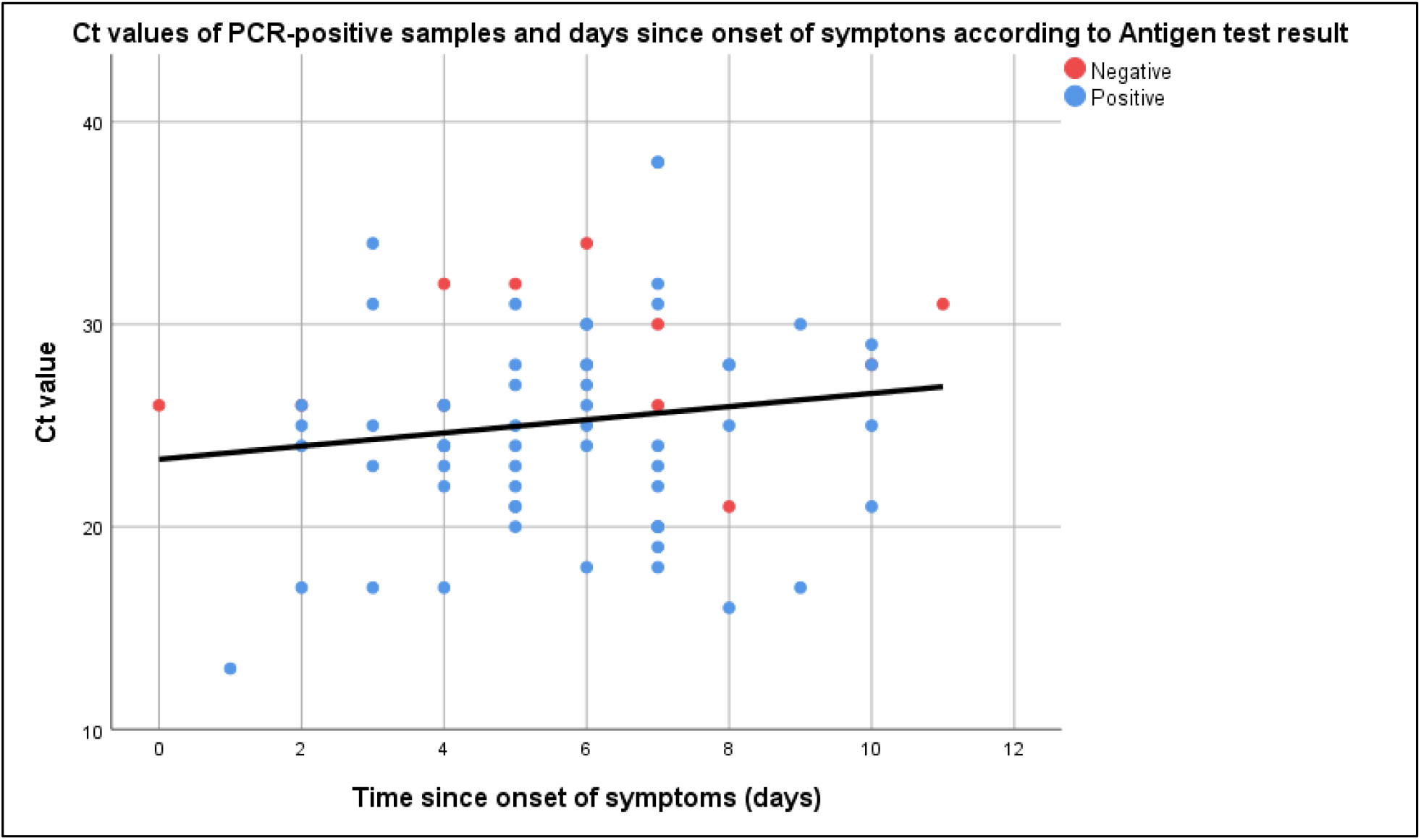
Ct Values of PCR-Positive Samples versus Days Post Onset of Symptoms.

## DISCUSSION

This study was designed to evaluate the performance of the Panbio™ COVID-19 Ag test in comparison with RT-PCR, considered the gold-standard diagnostic test for COVID-19. The overall sensitivity was 84% (95% CI, 75-93.8%) and the overall specificity was 98% (95% CI, 96-98.8%), similar to results seen in other recent studies^11,12^.

In our study, the concordance between the Panbio™ COVID-19 Ag test and RT-PCR decreases when Ct levels increase. In particular, the concordance was 97% below a Ct value of 25, but decreases at higher Cts. Additionally, sensitivity is not significantly different between subjects tested 0 to 11 days post symptom onset due to the high viral load observed in the patients even at 7 days. Following SARS CoV-2 infection, the virus undergoes a period of incubation during which viral titers are usually too low to detect, after which the virus undergoes exponential growth, leading to a peak in viral load and infectivity, and ending with declining viral levels and clearance when infectivity is low. For disease control, it is very important to identify infected individuals who present COVID-19 symptoms and also those who are suspected of infection due to contact with infected individuals. The Panbio™ COVID-19 Ag test has sufficient sensitivity and specificity to serve as a tool for the primary screening of such individuals. This is especially true in point-of-care and near patient settings where more elaborate laboratory facilities are lacking for RT-PCR.

## Data Availability

All data of the manuscript on

